# Neural Correlates of Behavioral Recovery Following Ultrasonic Thalamic Stimulation in Chronic Disorders of Consciousness

**DOI:** 10.1101/2023.07.13.23292523

**Authors:** Josh A. Cain, Norman M. Spivak, John P. Coetzee, Julia S. Crone, Micah A. Johnson, Evan S. Lutkenhoff, Courtney Real, Manuel Buitrago-Blanco, Paul M. Vespa, Caroline Schnakers, Martin M. Monti

## Abstract

**Background:** Treatments aimed at hastening recovery from disorders of consciousness (DOC; e.g., coma, the vegetative state) have lagged behind a rapidly advancing science of these conditions. In part, this is due to the difficulty in selectively targeting the many deep regions of the brain known to be key for recovery from DOC. The (re)emergence of low intensity focused ultrasound (LIFU) neuromodulation addresses this gap by providing a non-invasive, safe, and relatively low-cost means to exert neuromodulatory effects, anywhere in the brain, with relatively high spatial precision.

**Methods:** As part of this first-in-man clinical trial, a cohort of 10 patients with chronic DOC underwent two sessions of MR-guided thalamic LIFU, with concomitant functional neuroimaging, one week apart. Behavioral responsiveness, measured with the Coma Recovery Scale Revised (CRS-R), was assessed at multiple time-points both before and after each LIFU session. Changes in clinical score before and after each session were compared within subjects.

**Results:** This convenience sample of sample of chronic DOC patients included, at entry, 4 Minimally Conscious State plus (MCS+), 4 Minimally Conscious State minus (MCS-) and 2 Vegetative State (VS) patients (6 male; mean age = 39.1, mean time since injury = 56.75 months; 4 anoxic and 6 traumatic injuries). We find a significant linear increase over time in CRS-R total score with thalamic LIFU exposure. Functional imaging reveals changes in brain-wide activity and thalamo-cortical connectivity of the targeted thalamus (but not the contralateral, non-targeted, thalamus), during LIFU administration. Strikingly, these effects are associated with the degree of behavioral recovery observed following exposure.

**Discussion:** Collectively, these results are the first to suggest the efficacy of thalamic LIFU for the treatment of chronic DOC and extend our previous investigations in acute DOC populations. Indeed, results from both cohorts support the safety, feasibility, and preliminary efficacy of LIFU, as evaluated by gold-standard clinical assessments. Moreover, imaging results in both datasets provide a convergent biological link uniting neuromodulatory thalamic LIFU and the observed behavioral recovery. These first-in-man findings provide a key foundation to motivate further exploration of this technique (e.g., LIFU parameterization, optimal number and timing of exposures) and invite a sham-control clinical trial, in a larger cohort, to assess, in a blinded fashion, the technique’s efficacy.

Clinical Trial number, date of submission, date of first enrollment, registration link:

NCT02522429

August 13, 2015

March 10, 2016

https://clinicaltrials.gov/ct2/show/NCT02522429

## Introduction

Despite tremendous progress in understanding the mechanisms associated with loss and recovery of consciousness following brain injury^1^, there are still very few treatments available^2^ for the hundreds of thousands^3^ of patients, in the United States alone, who are in a Disorder of Consciousness (DOC)—such as coma, the Vegetative State (VS), and the Minimally Conscious State (MCS).^4^ One major obstacle in treating DOC, particularly in the context of device-based interventions, is the difficulty in accessing, in a non-invasive yet selective fashion, the deep structures of the brain particularly implicated in recovery of consciousness^1,5,6^. Indeed, multiple modes of invasive stimulation of the central thalamus (e.g., electrical^7– 9^, chemical^10^, and optogenetic^11,12^ stimulation of particular intralaminar nuclei—e.g., central lateral [CL] nucleus^7,8^) have shown to induce physiological and behavioral awakening in preclinical models, including rodents and non-human primates,^9,10^ during sleep and pharmacologically-induced unconsciousness. Consistent with these findings, deep brain stimulation of the central thalamus^7^ (CL) has shown the potential to elicit remarkable recovery in chronic DOC patients.^7,13^ Nonetheless, a 5-year prospective clinical trial concluded that a majority of DOC patients (∼87%) are unable to meet relatively minimal requirements for undergoing the surgery required for this procedure^14^—which, together with the known medical risks associated with surgical implantation,^15^ make this technique not broadly feasible despite its positive results. ^7,13^

Fortunately, recent years have seen the rapid maturation of Low Intensity Focused Ultrasound (LIFU)^16^, a technique capable of relatively selective neuromodulation to neural tissues anywhere in the brain^17^. Leveraging what is known of the neural correlates of DOC, this very first^18^ clinical use of LIFU aims to restore function in DOC patients through non-invasive central thalamic stimulation. Indeed, the current work is the first to provide the results of a full cohort of chronic (time since injury > 1year) DOC patients treated with this technique. Supported by the theory detailed above and our previously published works^19,20^, we hypothesize that DOC patients in the chronic phase of illness will demonstrate behavioral recovery following central thalamic LIFU. Importantly, chronic DOC patients^21^ are considerably more behaviorally stable when compared to acute patients^19^ and thus much less likely to recover spontaneously over the weeks observed here^21^, facilitating the attribution of improvements to our intervention.

## Methods

### Patients

This convenience sample included 10 chronic DOC patients (see Table 1; DOC at entry, 6 male, mean: age = 39.1, time since injury [TSI] = 56.75 months, 4/6 anoxic/traumatic injury) referred to the study by long-term care facilities and private households following the persistence of DOC past the 1-year post-injury period. An initial neurobehavioral evaluation with the JFK Coma Recovery Scale – Revised (CRS-R)^22^ was conducted prior to enrollment to confirm eligibility (i.e., a chronic DOC).

**Table 1).**
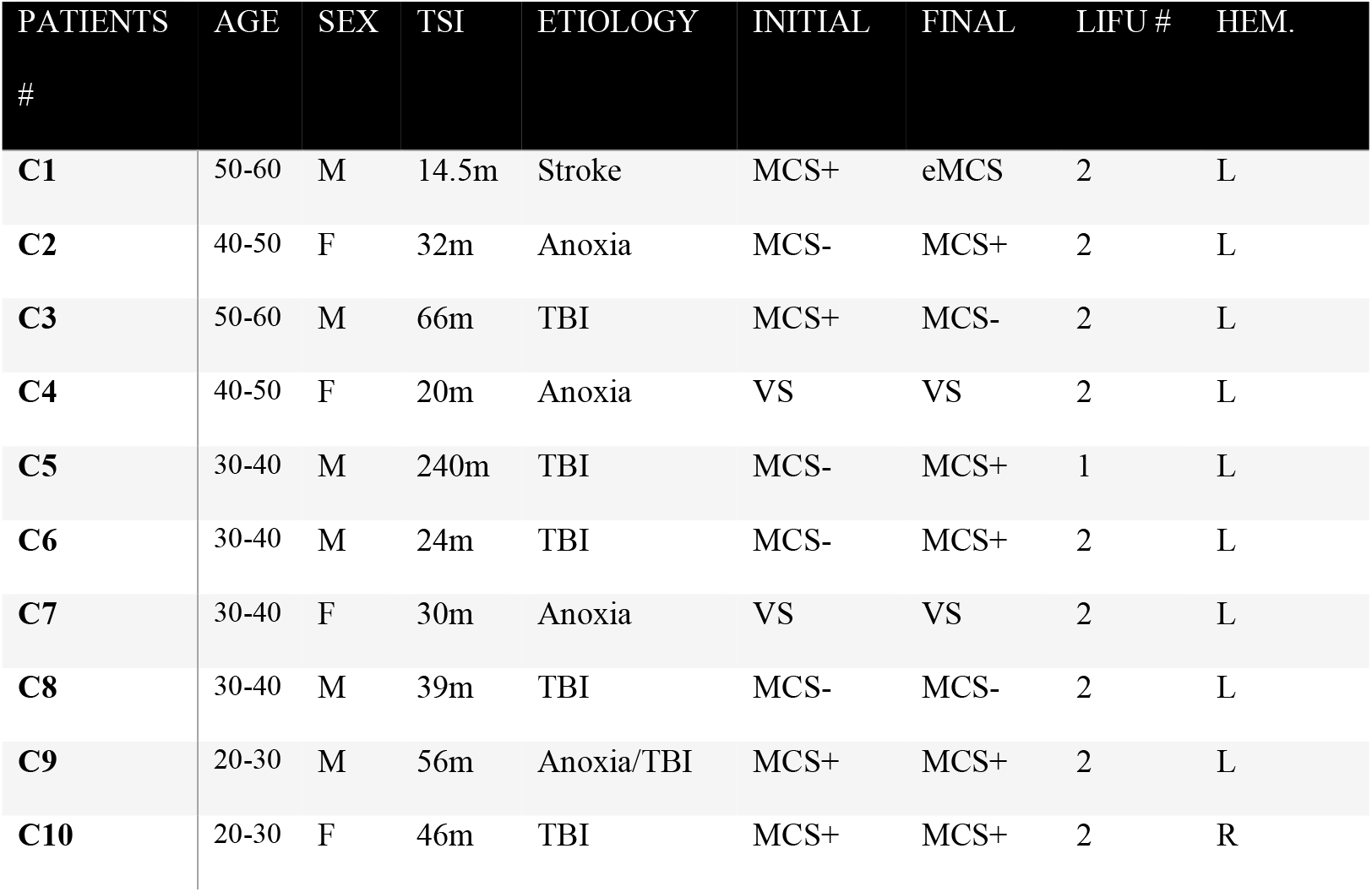
Relevant patient-specific information in subjects. TSI = time since injury. TBI = traumatic brian injury. MVA = motor vehicle accident. Initial diasnostic category (e.g., MCS+) is based on the highest performance measured using the CRS-R prior to LIFU exposure. Final diasnostic category is based on the highest performance measured using the CRS-R in the week following the final LIFU exposure (regardless if 1 or 2 sonications were performed). The number of sonications (LIFU #) and the hemisphere of the targeted thalamus (Hem.) is reported.

| PATIENTS<br># | AGE | SEX | TSI | ETIOLOGY | INITIAL | FINAL | LIFU # | HEM. |
| --- | --- | --- | --- | --- | --- | --- | --- | --- |
| C1 | 50-60 | M | 14.5m | Stroke | MCS+ | eMCS | 2 | L |
| C2 | 40-50 | F | 32m | Anoxia | MCS- | MCS+ | 2 | L |
| C3 | 50-60 | M | 66m | TBI | MCS+ | MCS- | 2 | L |
| C4 | 40-50 | F | 20m | Anoxia | VS | VS | 2 | L |
| C5 | 30-40 | M | 240m | TBI | MCS- | MCS+ | 1 | L |
| C6 | 30-40 | M | 24m | TBI | MCS- | MCS+ | 2 | L |
| C7 | 30-40 | F | 30m | Anoxia | VS | VS | 2 | L |
| C8 | 30-40 | M | 39m | TBI | MCS- | MCS- | 2 | L |
| C9 | 20-30 | M | 56m | Anoxia/TBI | MCS+ | MCS+ | 2 | L |
| C10 | 20-30 | F | 46m | TBI | MCS+ | MCS+ | 2 | R |

A diagnosis of DOC (specifically, VS or MCS) with a time since injury (TSI) greater than 1 year were the inclusion criteria. Deep pharmacological sedation, a history of neurological illness prior to injury, CRS-R results consistent with coma, and the inability to safely enter an MR environment (e.g., ferromagnetic non-MR safe implants) were exclusion criteria. Legal representatives of all patients consented to take part in the present study and to have the results of this research published.

### Experimental Design

The overall experimental protocol is shown in Figure 1A. Briefly, patients underwent at least three baseline neurobehavioral assessments (CRS-R^*22*^; at 1 week, 1 day and 1 hour prior to LIFU; henceforth, pre-LIFU) followed by a session of LIFU, and three additional neurobehavioral assessments (at 1h, 1 day, and 1 week following LIFU; henceforth, post-LIFU1). A second, identical, cycle of LIFU and assesssments were performed in all but one subject (C5; excluded due to infection). This included a second LIFU session (LIFU 2) 1-week following LIFU 1 with a behavioral assessment being done 1 hour, 1 day, 1 week, and 1 month following LIFU 2 (post-LIFU2).

**Figure 1).**
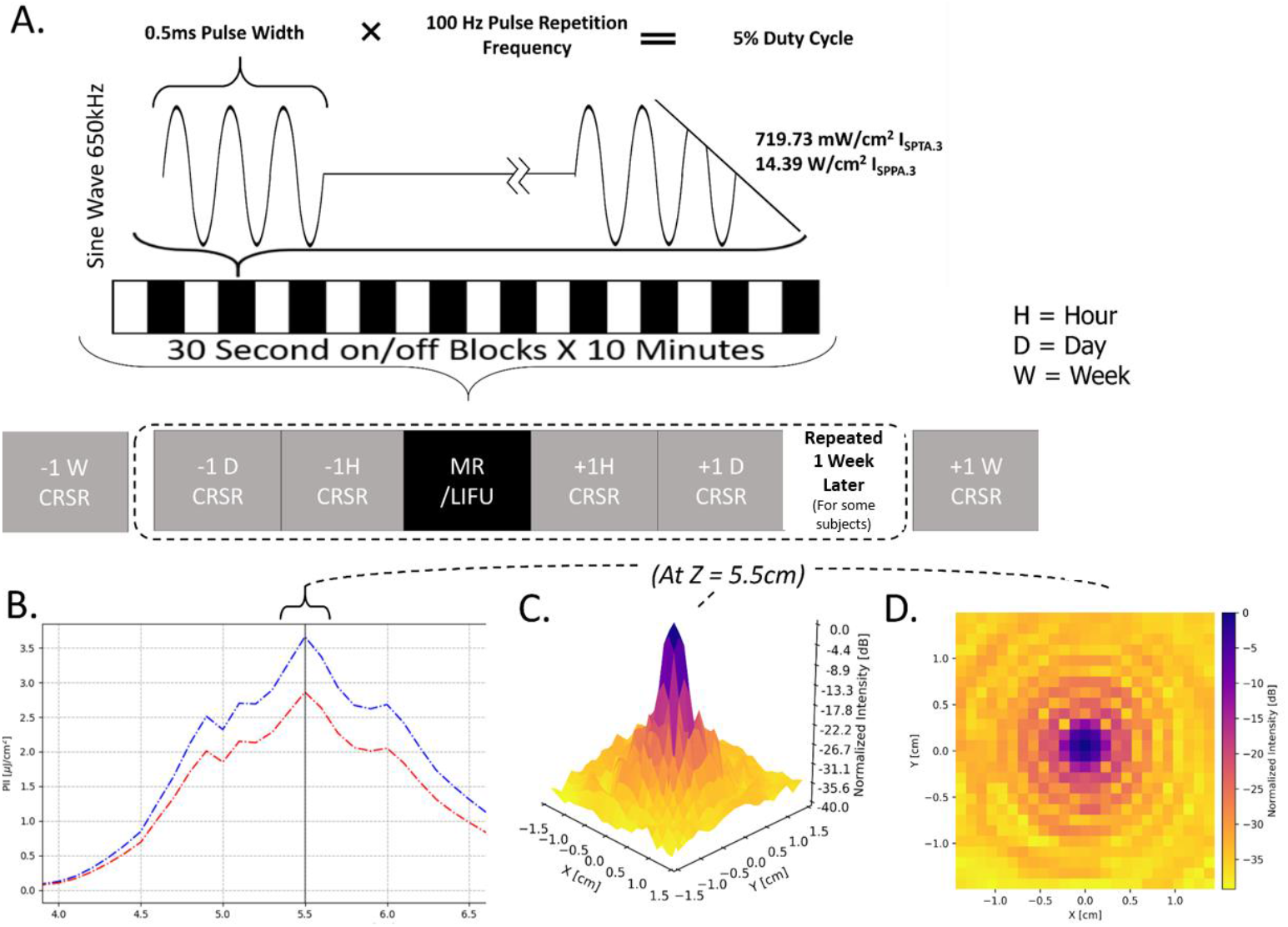
Depiction of study protocol involving LIFU parameters and CRS-R (Coma Recovery Scale – Revised) assessments. I_SPTA.3_ = Spatial Peak Temporal Average Intensity.3. I_SPPA.3_ = Spatial Peak Pulse Average Intensity.3 (“0.3” denotes deration (attenuation) due to absorption by tissue at 0.3 dB/cm-MHz). **1b)** Intensity (pulse intensity integral [PII]) in the longitudinal plane (Z plane, extending from transducer surface). Note the peak intensity 5.5 cm from the transducer surface and that a 50% (−3 dB) reduction in peak intensity is found in an area approximately 1.5 cm in length. **(1c**,**1d)** Intensity in the radial plane (X/Y plane, extending laterally from the focal point of ultrasound beam 5.5cm from transducer surface) shown in both 3 **(1c)** and 2 **(1d)** dimensions. From these graphs, note that a 50% (−3 dB) reduction in peak intensity is found in an area approximately 0.5 cm in width. This figure was first published in our previous work^19^, which employed a near-identical design in acute DOC patients.

### LIFU Sonication Protocol and Procedure

#### LIFU Sonication Parameters

In each session, LIFU was applied at 100 Hz pulse repetition frequency (PRF), 0.5ms pulse width (PW), 650 kHz carrier wave frequency, 5% duty cycle (DC), and 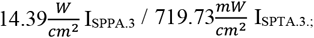 “.3” denotes tissue absorption at 0.3 dB/cm-MHz. This parameter set (PRF/PW/DC) has been derived from prior work demonstrating its neuroactivity^*23,24*^. In regards to safety, these energy levels fall below the FDA limit for diagnostic ultrasound imaging of the human cranium^*25*^ and are far below those demonstrated to avoid inducing histological damage^*26*^. The LIFU waveform was emitted from a single-element transducer (Brainsonix; 71.5mm curvature) positioned, using MR-guidance, such that its theoretical focus (55mm from its surface) lay over the intended thalamic target. Once appropriate transducer placement was confirmed visually (see below), ultrasound was delivered inside the MRI for a total of ten 30-second on blocks, separated by 30s off periods (see Figure 1).

#### LIFU Target

In light of the results from thalamic deep brain stimuation (DBS) in DOC^*7,14*^ and prior theoretical^*6*^ and empirical^*5,19,20,23,27*^ work concerning the role of the thalamus in these conditions, the intended LIFU target was the central thalamus. More specifically, we attempted to target the caudal region of the CL nucleus as well as the centromedian (CM)–parafascicular (PF) nuclear complex, though much of the central thalamus and adjacent thalamic tissue were certainly also directly influenced—especially when considering the substantial thalamic atrophy in our cohort. The protocol called for sonication to occur preferentially to the left thalamus, on the basis of prior work suggesting a generally greater link between left thalamic damage and DOC symptoms^*5,27*^. Nonetheless, the right thalamus was chosen in patients with left craniectomy, in order to avoid the possibility of higher than expected energy deposition given the unknown penetration and refraction profile of ultrasound passed through synthetic bone replacement materials. The same precaution was excercised in the case of implanted medical devices (e.g., stints, ventricular shunts) proximal to the intended left thalamic target and potentially resulting in risk to the patient given the possibility for unknown interactions between these devices and the applied ultrasound (See Table 1 for the hemisphere of LIFU administration in this cohort.)

#### LIFU Procedure

The area surrounding the planned LIFU entry point on the head was shaved prior to positioning in order to minimize the possibility of trapped air bubbles under the transducer and, thus, the impedence of ultrasound. Just prior to scanning, ultrasound gel (aquasonic) was applied to this region and smoothed in order to remove any air pockets. A thin layer of gel was applied to the surface of the transducer and any bubbles, if present, were similarly smoothed from this layer. The transducer was then coupled to the head with gel filling any open space between the transducer face and the scalp. The ultrasound transducer was then positioned so that its center lay over the squamous portion of the temporal bone (generally, the thinnest part of the human skull) in a perpendicular trajectory through the skull and into the thalamus, in order to minimize bone-induced ultrasound attenuation and refraction^*28*^. Next, two straps— one horizonal and one vertical—were used to secure the device to the patient. Conventional soft foam padding and pillows were used to further secure the positioning of the device and decrease the potential for head motion during the procedure. Next, we acquired a rapid (95 s) T1-weighted MPRAGE anatomical image (see “MRI Sequences” for sequence). Using a circular MR fiducial, the visible center of the transducer, and the known focal depth of our transducer, reference lines were drawn with the Siemens 3D display GUI which depicted the theoretic focus of the transducer. Adjustments to the positioning of the transducer on the head were made iteratively, re-acquiring a T1-weighted MPRAGE at each iteration, until the beam trajectory from the center of the transducer was assessed to be in-line with the intended thalamic target.

### Safety Measures

With respect to safety, we recorded vital parameters during LIFU administration and MR data collection (e.g., heart rate, blood oxygen, blood pressure). Furthermore, any adverse events that occurred to patients over the course of the study were documented through medical records and reviewed by a neurointensive care specialist (PV).

### Behavioral Analysis

Behavioral data analysis was performed using JASP (JASP Team (2019). JASP, Version 0.11.1).

Behavioral responsiveness in patients was assessed using the total score on the CRS-R^*22*^ as well as CRS-R^_index*29*_^, which is derived from the raw CRS-R score using R (Rstudio2021; https://github.com/Annen/CRS-R/blob/master/CRS-R_index.R). The CRS-R_index_ is a single value calculated from CRS-R subscores and was chosen because it is thought to more appropriately represent functional recovery with a single number.

Behavioral statistics were done by grouping the highest CRS-R or CRS-R_index_ score for each experimental period (i.e., Pre-LIFU 1, Post-LIFU 1, and Post-LIFU 2) in order to best capture patients’ maximal performance. Because C5 did not receive two sessions, this subject was excluded in our behavioral analyses. A repeated measures ANOVA was run to assess a main effect of time-point on responsivness. For both the CRS-R and CRS-R_index_ data, Mauchly’s Test of Sphericity found that the assumption of sphericity was met for both datasets.

Furthermore, as a linear trend appeared likely when plotting each time-point, this was assessed for statistical significance.

For inclusion in MRI data analysis, a single “recovery” value was created by subtracting the highest Pre-LIFU score from the highest Post-LIFU 2 score, which was used as a regressor in group-wide analyses (see details below). In this case, only the CRS-R_index_ was used because its non-ordinal data are more appropriate for inclusion as a regressor in MRI group statistics.

### MRI analysis

Complete details concerning MRI analysis are presented in the supplemental methods section. However, in brief: during each session, T1-weighted MPRAGE structural images were taken prior to LIFU while blood oxygen level dependent (BOLD) functional images were taken during LIFU administration. Volumes which coincided with the onset and offset of 30s-long LIFU on/off blocks were recorded. Additionally, in a subset of patients, functional resting-state images were taken both prior to and following LIFU administration.

All functional images were preprocessed in FSL (FMRIB Software Library v6.0.1)^*30*^ using methods which correspond to those recommended in a recent empirical analysis of appropriate preprocessing methods for DOC patient MRI data^*31*^ and subsequently used in the following analyses: **1)** BOLD signal throughout the brain during LIFU-on and LIFU-off blocks was contrasted within subjects in order to determine if activity in any region was acutely altered by LIFU. These results were aggregated at the group level in a fixed-effects model and cluster-corrected at z > 3.09 (p < 0.05) and, separately, at z > 2.56 (p < 0.05), which is appropriate for FS^*32*^; group-level statistics will mirror these for all analyses of functional data. Morevoer, in a separate model, the degree to which subjects recovered (difference in maximal CRSR_index_ at baseline and following final sonication) was used as a regressor at the group level to assess if and where changes in BOLD corresponded to behavioral outcome. **2)** Spherical regions of interest (ROI) were created around each thalami; these were used to extract mean z-scores for each subject corresponding to the degree to which LIFU modulated BOLD activity specifically within each thalamic ROI (using whole-brain z-score maps produced in analysis 1). JASP (Version 0.11.1) was used to statistically assess if these values for the target thalamus differed from zero (null effect), differed from the non-target thalamus, or correlated with behavioral recovery. **3)** The time-course of BOLD signal within each thalamic ROI was also extracted for each subject/session and used to perform a psycho-physiological-interaction (PPI)^*33*^ analysis, which assessed if connectivity between each thalamus and the rest of the brain changed during LIFU-on blocks compared to baseline. Again, a separate model at the group-level was used to determine if these PPI results were modulated by the degree of behavioral recovery and, if so, where. **4)** Finally, in a supplemental analysis, connectivity between each thalamus and the rest of the brain was calculated for resting state images taken prior to and following LIFU for those subjects possessing that data. The resultant connectivity maps were subtracted within subjects and aggregated at the group level to determine if this connectivity changed following LIFU.

## Results

### Safety

One adverse event (AE) was recorded throughout the entirety of the study. Specifically, one patient contracted a respiratory infection during the course of the study. However, clinical review deemed this AE to be unrelated to LIFU administration. Similarly, no alteration of vital parameters was observed during LIFU administration.

### Behavioral Analysis

Analysis of the highest CRS-R_index_ score within each period (i.e., baseline, post-LIFU1, post-LIFU2) revealed a significant positive linear trend (p=0.040; see Figure 2), with maximum responsiveness progressively increasing after thalamic sonication. The same significant trend was also observed when analyzing the highest CRS-R total score for each period (p = 0.019). In either analysis, no significant associations were found between change in responsiveness and age, TSI, etiology, or baseline responsiveness (i.e., highest CRS-R_index_ or CRS-R total score at baseline). Furthermore, no correlations between recovery and baseline responsiveness were found, in contrast to our prior work in acute DOC patients^**19**^. Importantly, in 4 of the 10 subjects, the observed gains in responsiveness represented a shift upward in diagnostic category (e.g., see Table 1). However, the reverse pattern was observed in one patient, with clinical responsiveness decreasing after LIFU, and returning to baseline levels at the 1-month follow-up.

**Figure 2).**
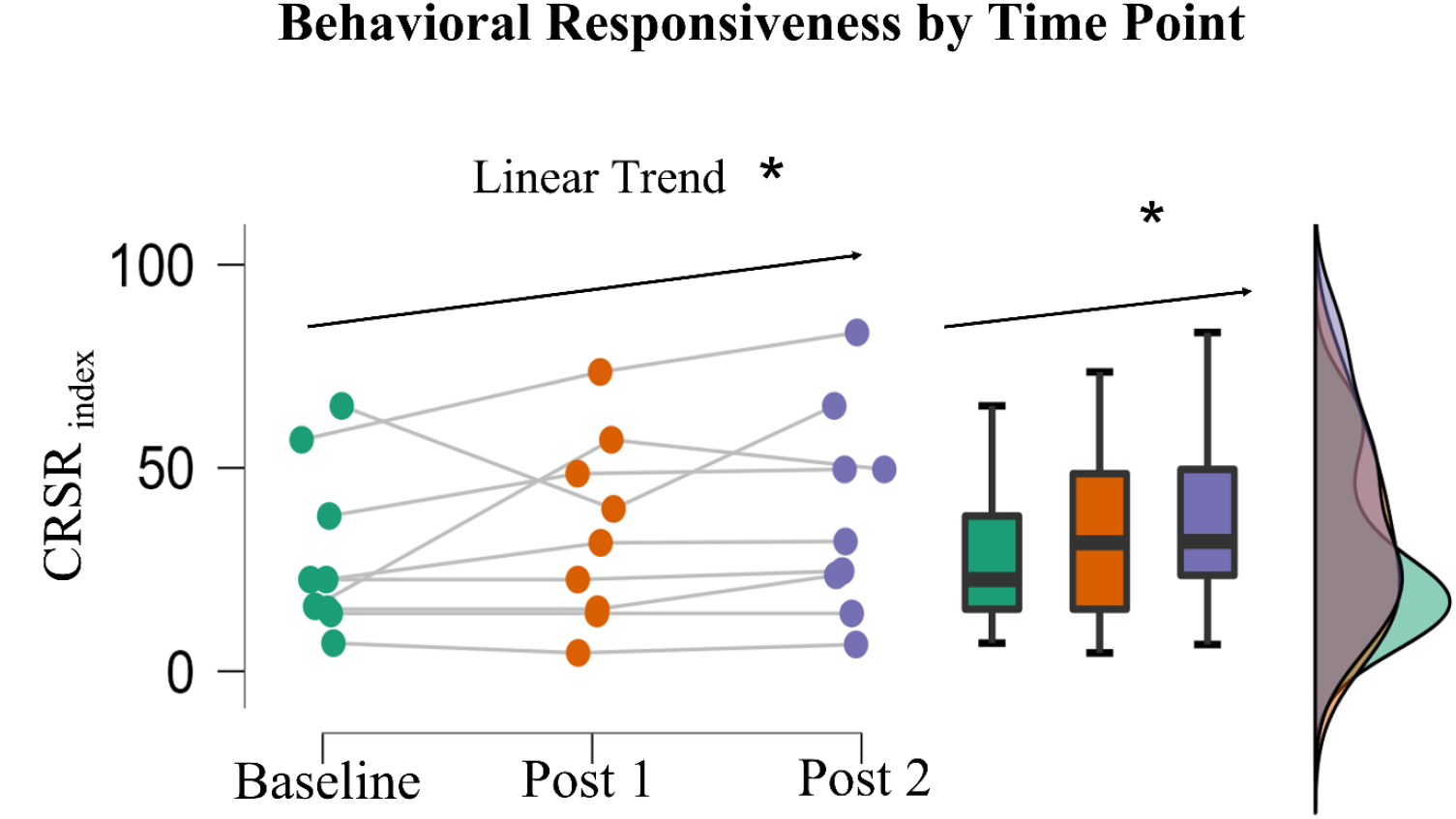
Cluster, box, and raincloud plots display the distribution of the highest CRS-R_index_ score prior to and up to 1-week following LIFU 1 (Post 1) and LIFU 2 (Post 2). Bold solid lines in boxplots represent the mean of each distribution while upper and lower bounds of each box represent the first and third quartile. A significant positive linear trend was found.

### MRI analysis

#### BOLD Data Analysis: Effect of LIFU on Activity

As compared to baseline (i.e., LIFU-off), 30 second blocks of deep brain LIFU sonication coocurred with significantly reduced BOLD signal in one temporal cluster (see Figure 3a), ipsilateral to the site of stimulation. However, behavioral recovery was associated with a reduction in BOLD signal within one cluster located in the ispilateral subcortex, which appears to specifically surround the anterior ventral striatum in most subjects (see supplementary figures S1).

**Figure 3).**
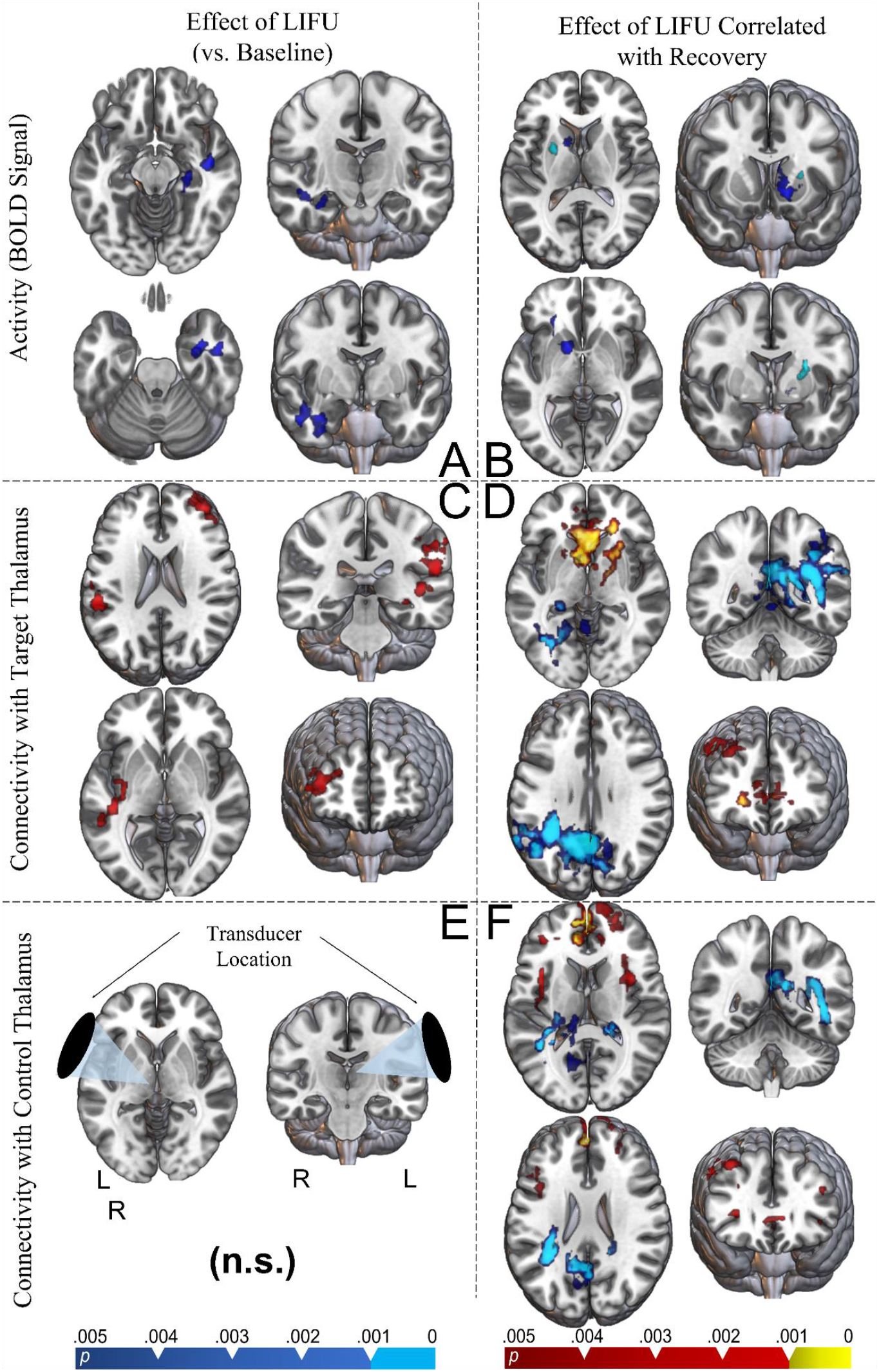
Whole brain results in standard (MNI) space. For all analyses, statistical maps were obtained using a fixed effects model as implemented in FSL 6.0.1, and are shown at two levels of cluster correction for multiplicity. For inhibition/reduction,a cluster-defining threshold (CDT) set at p < 0.005 is in blue and CDT at p < 0.001 is in violet. For activation/enhancement, a CDT set at p < 0.005 is in red and CDT at p < 0.001 is in yellow. **A)** Regions of significant change in BOLD signal during sonication compared to inter-sonication periods (i.e., baseline). **B)** Regions of significant BOLD change predicted by behavioral recovery. **C)** Connectivity changes observed during LIFU-on blocks compared to LIFU-off blocks (PPI) between the whole brain and the targeted thalamus. **D)** PPI changes (target thalamus) predicted by behavioral recovery. **E)** Connectivity changes observed during LIFU-on blocks compared to LIFU-off blocks (PPI) between the whole brain and the non-targeted (control) thalamus. **F)**PPI changes (non-target thalamus) predicted by behavioral recovery. Note that our patients often had brains that deviate signficiantly from the healthy brains used to create the MNI atlas. Thus, see the same results projected onto a group-wide composit T1-weighted image in supplementary figure S2.

#### BOLD Data Analysis: Connectivity

Psycho-physiologiocal interaction (PPI) analysis found that, during LIFU sonication, the connectivity of the targeted thalamus increased with three clusters—one extending from the ipsilateral opercular cortex to the ipsilateral supramarginal gyrus, one in the contralateral frontal polar cortex, and one in the ipsilateral insular cortex (Figure 3c). Conversely, when the same analysis was performed on the non-targeted (i.e., contralateral) thalamus, no significant change in connectivity was found during sonication.

Furthermore, the pattern of increased and decreased connectivity of the targeted thalamus during LIFU was found to be associated with the degree of recovery observed following the intervention (i.e., change in CRS-R_index_), potentially providing a mechanistic link between sonication and the ensuing recovery. Specifically, increased recovery was associated with increased connectivity between the targeted thalamus and a cluster spanning the contralateral striatum, globus pallidus, and anterior cingulate (cluster 1) as well as a cluster in the contralateral middle frontal gyrus (cluster 2). Increased responsiveness following LIFU was also observed to be associated with decreased connectivity between the targeted thalamus and the posterior cingulate cortex during sonication (Figure 3d).

Although, as mentioned above, the non-targeted thalamus did not exhibit changes in connectivity as a function of whether LIFU was being administered (Figure 3e), it did exhibit an association between changes in connectivity during sonication and the degree of ensuing recovery (i.e., change in CRS-R_index_) that mostly mirrored what was observed in the targeted thalamus (Figure 3f).

#### BOLD Data Analysis: Connectivity (Pre – Post)

When comparing resting-state BOLD signal collected (minutes) prior to and following LIFU, longer-term changes in connectivity between both thalami (targeted and non-targeted) and the rest of the brain were found, which are depicted in supplementary figures S3-4. Though these results are difficult to interpret, given the extensive number of signficant clusters found, in general, connectivity between the targeted thalamus appears to decrease with large portions of frontoparietal cortex following LIFU, while connectivity changes between the control (non-target) thalamus and the rest of the brain appear more mixed—including an increase in connectivity with many portions of frontoparietal cortex. Notably, both thalami appear to show persistent decreases in connectivity with the midline and ipsilateral posterior cortical regions whose decreased connectivity with the target thalamus appears to predict recovery (see Figure 3d) in these subjects as well as with the temporal region shown to reduce in BOLD signal during LIFU blocks (see Figure 3a).

## Discussion

This study collectively joins our previously-published^**19**,**20**,**34**^ parallel works as the first clinical application of non-destructive focused ultrasound neuromodulation. Despite the pragmatic limitations of this first-in-man exploration—which are discussed more below and in prior work^**19**,**20**^—these results collectively suggest the safety, feasibility and initial efficacy of thalamic LIFU as an intervention in disorders of consciousness while also providing a first glimpse at its neural mechanisms of action.

Firstly, with respect to feasibility and safety, our results mirror previous findings^**19**,**20**,**34**^ which support the safety of thalamic LIFU, at these parameters. Indeed, we only observed a single adverse event, which was determined to be independent of the sonication itself, and, across 19 LIFU administrations, we observed no alteration of vital parameters. Moreover, these results show that MR-guided thalamic LIFU is feasible in this patient population and can produce viable MRI data, despite the known difficulties imposed on MRI analysis by severe brain injury^**31**^.

Secondly, with respect to behavior, we observed a linear increase in the neurobehavioral responsiveness of this cohort following each of two thalamic LIFU sessions. This is in contrast with the possibility of a “leveling out” of recovery following the first LIFU; although these results should be considered preliminary, they may suggest a dose-response effect whereby multiple LIFU sonication sessions could provide additional benefits to patients. For 4 of 10 subjects, behavioral changes included a shift up in diagnostic category (see Table 1), which implies the emergence of novel behaviors not observed prior to ultrasound administration and which might positively affect quality of life for a patient (e.g., response to command, communication). These observed behavioral changes are particularly compelling considering that, in contrast to acute DOC,^**19**,**34**^ these chronic patients are more behaviorally stable and are much less likely to recovery spontaneously—especially in the time frame of weeks they were observed for in this study^**21**^.

Finally, significant fMRI results provide preliminary data concerning a mechanistic link between sonication and ensuing behavioral changes. Paralleling previous results in acute patients^**20**^ and in healthy individuals^**24**^, we find reduced BOLD signal during thalamic LIFU sonication (i.e., LIFU-on), compared to periods of no sonication (i.e., LIFU-off), in cortical regions remote to the site of stimulation. We do note, however, that the localization of group-wide results in the temporal lobe may be unexpected given what has been reported in other instances of thalamic LIFU and may be due to multiple limitations, as discussed below. Yet, and despite differences in the precise pattern of effects, reduced BOLD signal during LIFU blocks using the precise parameters employed here (e.g., 5% DC, 100Hz PRF, etc.), has now been shown across three separate cohorts^**19**,**24**^.

Perhaps more compellingly, inhibition of subcortical tissues ipsilateral to the site of stimulation was associated with recovery in chronic subjects (See Figure 3b). In standard space, this (in)activation appears to reside within the ipsilateral striatum, which is further supported by projecting this group-wide cluster on the brains of single subject (see Figure 3b, Figure S3). Broadly, reduced striatal output has been hypothesized as a mechanism underlying DOC^**6**^, while, specifically, the ventral striatum is a component of the basal forebrain nuclei, which share intimate reciprocal thalamic connectivity and play a key role in maintaining cortical arousal^**35**^. These results may conceivably stem from two sources: **1)** Direct inhibition of the region during sonication, given its proximity to thalamus or **2)** circuit-level effects during sonication. The first hypothesis of direct stimulation appears unlikely given the distance (> 2.5cm) of this cluster from the central thalamus and generally far lower previous estimates^**24**,**36**^ and empirical examples^**37**^ of skull-refractory effects on focal location in comparable conditions. Conversely, the second hypothesis—the downstream inhibition of the striatum following thalamic inhibition—seems plausible considering the intimate relationship between thalamic output and striatal input from the cortex. The established relationship between these regions is such that reduced thalamic output (consistent with the notion of local inhibition of the thalamus^**24**,**38**^) is likely to co-occur with reduced striatal activity; indeed, such a relationship is thought to underlie some aspects of the broad reduction in cortical activity observed in DOC patients^**6**^. While the observation that reduced striatal activity—a correlate of DOC—appears to predict recovery in these patients may appear counterintuitive, it should be noted that we have no data from which to understand the longer-term activity of this or any other of the relevant brain regions. The analyses reported only suggest that reduced activity in some regions during 30-second-long LIFU-on compared to LIFU-off blocks may be important for recovery. It is indeed conceivable that recovery could be driven by increased activity in these regions as a rebound from the immediate effect of LIFU rather than a reduction in activity compared to a pre-LIFU baseline. While our data cannot address this hypothesis, future studies should include more extended pre-LIFU baselines in order to clear up this ambiguity, as discussed more below.

The above results are perhaps better contextualized by our connectivity (PPI) results. During LIFU-on blocks, compared to LIFU-off blocks, the targeted thalamus increased its connectivity with several cortical clusters. Given that DOC is often hypothesized as a “disconnection syndrome”, where reduced functional communication (especially between subcortical structures and association cortex) underlies dysfunction^**6**^, it is interesting to observe enhancement of functional connectivity between thalamus and association cortex during LIFU in these subjects. Yet, the relationship between changes in connectivity and recovery appears more complex. We observe recovery-related increases in connectivity between thalamus and the bilateral frontal cortex as well as recovery-related reductions in connectivity between thalamus and the PCC and ipsilateral parietal cortex during LIFU. This general pattern was observed for both the targeted and non-targeted thalamus; however, changes in connectivity with the non-targeted thalamus appear notably less expansive. The possibility that the natural correlation between each thalamus might underlie this overlap should be considered. The presence of recovery-related changes in connectivity with the targeted thalamus with large sections of association cortex should likely be emphasized over the specific regions in which those changes are found, especially considering the broad connectivity of the thalamus. However, the observation of recovery-related changes in connectivity between thalamus and the PCC, lateral parietal cortex, and (especially medial) frontal cortex is very relevant when considering their inclusion in the classical default mode network—dysregulation of which has previous been observed in DOC patients ^**39**^. Moreover, long-term changes in connectivity were found between both the targeted and non-targeted thalami and the rest of the brain following LIFU in a subset of our cohort in which offline data was collected (see figures S3-4). These findings included both increases and decreases in connectivity, which is consistent with the view that recovery from DOC is paralleled by a restoration of appropriate patterns of both positive and negative connectivity across brain-wide networks^**39**^. Nonetheless, considering the fact that this is a small sample by functional imaging standards and that detailed interpretation of activations should be cautious, these results do suggest that longer-term changes in network structure appear to occur following LIFU in this cohort, something that may explain how our behavioral results appear to lag behind stimulation, but also last for a long period following stimulation.

### Synthesis of Results with Previous Findings

While some clear differences exist between near-identical analyses performed on a previously reported acute cohort^**19**^ and the present chronic cohort (e.g., the precise pattern of fMRI effects found), several overarching observations can be emphasized when considering the results in the aggregate. First, behavioral responsiveness was observed to increase following LIFU, on a group-wide level, in both cohorts. Second, changes in connectivity were observed between the targeted thalamus and cortical regions in both cohorts while no changes were observed between the non-targeted (control) thalamus and the rest of the brain. While the precise pattern observed differed in acute and chronic patients, both included changes in connectivity between thalamus and striatum as well as frontal cortical regions, which are known to be connected with the intended (central) thalamic target^**7**,**40**^. Finally, changes in connectivity between the targeted thalamus and cortex were associated with the ensuing recovery in both cohorts, which jointly link the immediate effects of thalamic sonication on neural activity with the clinical improvements observed in the following days.

One interesting aspect of our collective results is that, in parallel with work in healthy volunteers,^**24**^ sonication with our specific parametrization appears to be associated mainly with reduced BOLD signal during LIFU. At least at face value, it might appear counterintuitive that increased responsiveness follows from acutely decreased metabolic response, as inferred with BOLD imaging. Indeed, the mesocircuit hypothesis–the dominant framework for understanding impairment and recovery in disorders of consciousness–focuses on reversing a pathological depression in thalamic glutamatergic output^**6**^. Nonetheless, preclinical work in the rodent model shows that thalamic inhibition results in decreased inhibitory currents at the cortical level^**41**^; this “disinhibition” could, itself, lead to the observed decrease in BOLD signal—either by reducing the high metabolic cost^**42**,**43**^ and/or the vasodilatory effect of specific interneuron populations^**44**^ or by decreasing the energetic cost of action potentials in excitatory neurons once interneurons are silenced^**45**,**46**^ (i.e., during sonication). Thus, as discussed elsewhere,^**19**,**47**^ particularly in patients with low levels of basal neural metabolism,^**48**^ inhibitory perturbation of thalamus could help shift the thalamocortical system away from the pathological local minima in its functional state space that are thought to entrench some DOC patients in a state of disfunction.^**1**^ Supporting this, some CNS depressants (e.g., zolpidem, baclofen, lamotrigine, lorazepam) have been associated with recovery in select DOC patients^**49**,**50**^ in a way that is both sustained and apparently unrelated to the pharmacokinetics of the employed drugs^**47**^, adding credence to the idea that latent functional states may be accessed through inhibitory perturbation.

As mentioned above, there also are several differences in the patterns of functional changes observed in the present cohort, compared to the previously reported acute one, despite almost identical methodology.^**19**^ A number of different causes could underlie the discrepancies. First, the two cohorts are at very different time-points in the pathophysiology of brain injury^**51–54**^, which could result in different neural and/or vascular response profiles to thalamic sonication. Similarly, the known patterns of brain tissue atrophy typical of DOC patients^**5**,**55**^ are expressed over time and affect the very subcortical regions (e.g., thalamus, basal ganglia) targeted by our intervention. Finally, as discussed further in the limitation section, it has to be acknowledged that the specific brain topology of fMRI studies is less reliable in small sample sizes^**56**^, something which is further compounded by the known difficulty in acquiring^**31**^ and processing^**57**^ this kind of datum in the presence of severe pathology.

## Limitations

This work necessarily suffers from a number of limitations inherent to early phase clinical trials, to the characteristic complexities specific to studying DOC patients, as well as those introduced by our stimulation method. First, while the present (and prior) work was meant as a first-in-human trial, it does invite a double-blind sham-controlled trial in a larger population to appropriately estimate the effect size of thalamic LIFU in this disease^**58**^. Second, the high degree of variability in brain morphology in chronic DOC subjects likely biases our MRI results to effects which are spatially expansive and thus more detectable even in the presence of imperfections in data alignment (i.e., registration) for group analysis. Third, more timepoints involving the collection of biomarker data would further clarify the mechanisms linking sonication to the observed clinical amelioration. Currently, we can only speculate on how the functional changes observed during LIFU administration develop in the fullness of time and relate to more prolonged changes in behavioral responsiveness. Finally, it is well-known that a degree of attenuation and refraction of the focal point of LIFU is observed when passing through the human skull, and that this can vary greatly between individuals due to different skull shapes and densities^**28**^. Technologies which use multiple transducer elements to measure skull-related attenuation and refraction of LIFU and account for these effects are currently in development^**59**^. Future studies would benefit greatly from this capacity by ensuring more precise targeting of (even individual) thalamic nuclei and that sufficient, yet safe, energy is applied to the target. Similarly, recent data on the safety of LIFU suggests that the upper-limit of safe in-brain intensities is likely far higher than that applied here^**26**^, opening the door to investigating dose response effects at higher LIFU intensities.

## Conclusion

Collectively, the work presented here and our prior report in acute DOC patients suggest that thalamic LIFU is a feasible and safe intervention in this patient cohort. While preliminary, our results also show initial signs of effectiveness, which is particularly remarkable in chronic patients for whom spontaneous recovery is unexpected over such short periods of time. Finally, we also provide a potential biological link uniting acute exposure to thalamic LIFU and its effects on thalamocortical interactions with the clinical improvements observed over the following days.

## Supporting information

Supplemental Methods and Results

## Data Availability

Anonymized data not published within this article can be requested through a Material Transfer Agreement (MTA) with the UCLA TDG office.

