## Supplemental Methods and Results for "Neural Correlates of Behavioral Recovery Following Ultrasonic Thalamic Stimulation in Chronic Disorders of Consciousness"

‡ Joint last authors

#### Abstract

Background

Treatments aimed at hastening recovery from disorders of consciousness (DOC; e.g., coma, the vegetative state) have lagged behind a rapidly advancing science of these conditions. In part, this is due to the difficulty in selectively targeting the many deep regions of the brain known to be key for recovery from DOC. The (re)emergence of low intensity focused ultrasound (LIFU) neuromodulation addresses this gap by providing a non-invasive, safe, and relatively low-cost means to exert neuromodulatory effects, anywhere in the brain, with relatively high spatial precision.

Methods

As part of this first-in-man clinical trial, a cohort of 10 patients with chronic DOC underwent two sessions of MR-guided thalamic LIFU, with concomitant functional neuroimaging, one week apart. Behavioral responsiveness, measured with the Coma Recovery Scale Revised (CRS-R), was assessed at multiple time-points both before and after each LIFU session. Changes in clinical score before and after each session were compared within subjects.

Results

This convenience sample of sample of chronic DOC patients included, at entry, 4 Minimally Conscious State plus (MCS+), 4 Minimally Conscious State minus (MCS-) and 2 Vegetative State (VS) patients (6 male; mean age = 39.1, mean time since injury = 56.75 months; 4 anoxic and 6 traumatic injuries). We find a significant linear increase over time in CRS-R total score with thalamic LIFU exposure. Functional imaging reveals changes in brain-wide activity and thalamo-cortical connectivity of the targeted thalamus (but not the contralateral, non-targeted, thalamus), during LIFU administration. Strikingly, these effects are associated with the degree of behavioral recovery observed following exposure.

Discussion

Collectively, these results are the first to suggest the efficacy of thalamic LIFU for the treatment of chronic DOC in a full cohort of patients and extend our previous investigations in acute DOC populations. Indeed, results from both cohorts support the safety, feasibility, and preliminary efficacy of LIFU, as evaluated by gold-standard clinical assessments. Moreover, imaging results in both datasets provide a convergent biological link uniting neuromodulatory thalamic LIFU and the observed behavioral recovery. These first-in-man findings provide a key foundation to motivate further exploration of this technique (e.g., LIFU parameterization, optimal number and timing of exposures) and invite a sham-control clinical trial, in a larger cohort, to assess, in a blinded fashion, the technique's efficacy.



Clinical Trial number, date of submission, date of first enrollment, registration link:
NCT02522429
August 13, 2015
March 10, 2016
https://clinicaltrials.gov/ct2/show/NCT02522429

#### Supplementary Results

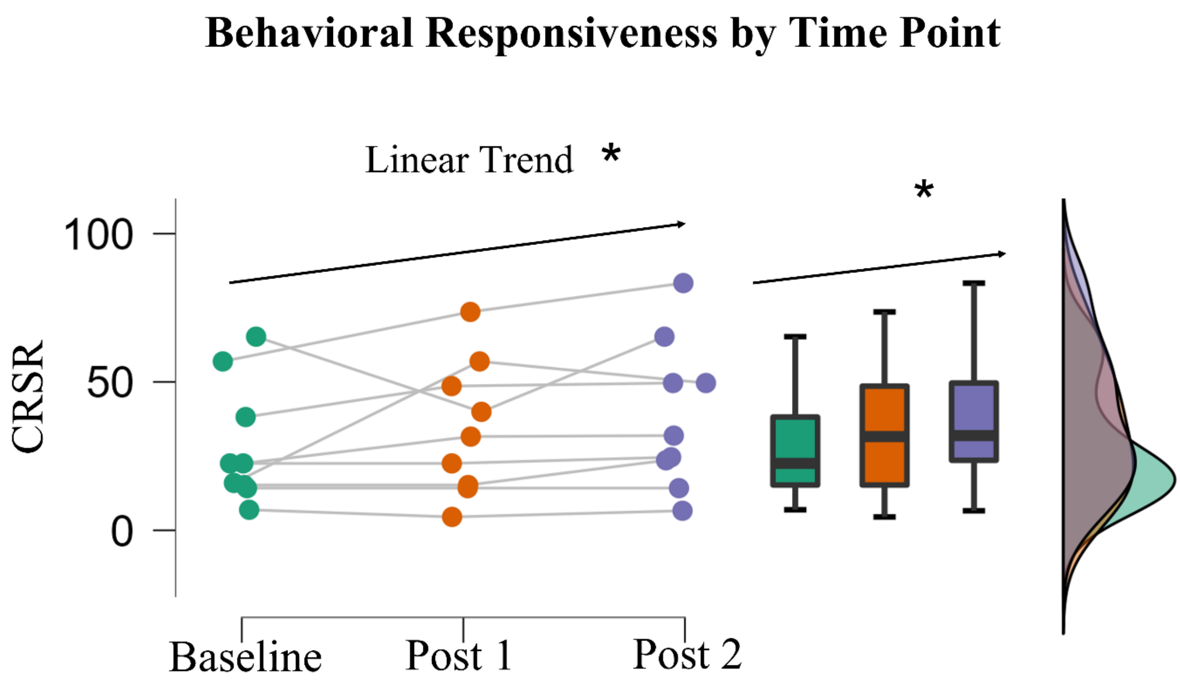


**Figure S1)** Cluster, box, and raincloud plots display the distribution of the highest CRS-R score prior to and up to 1-week following LIFU 1 (Post 1) and LIFU 2 (Post 2). Bold solid lines in boxplots represent the mean of each distribution while upper and lower bounds of each box represent the first and third quartile. A significant positive linear trend was found.

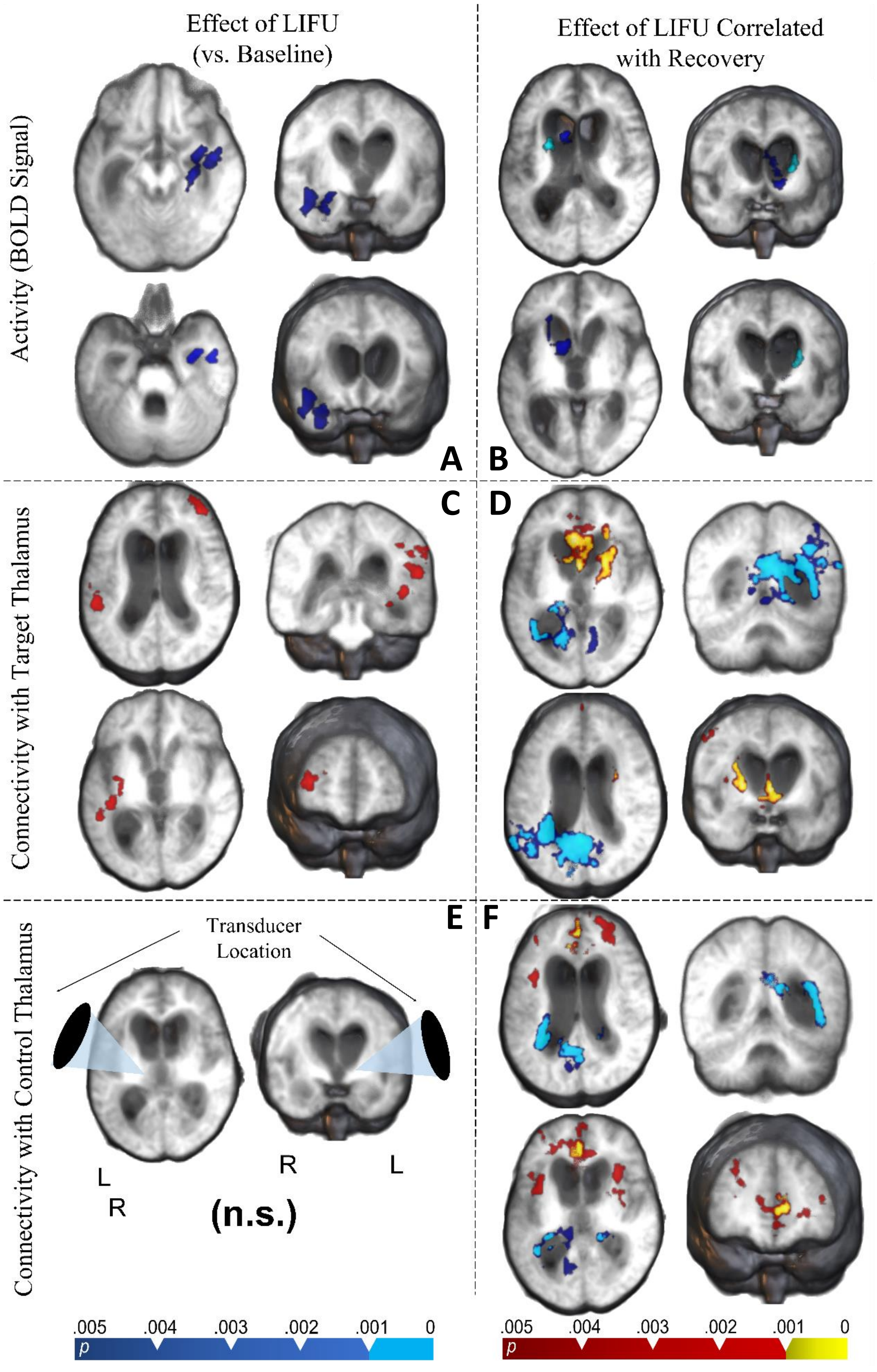

Effect of LIFU
(vs. Baseline)
Effect of LIFU Correlated
with Recovery
Activity (BOLD Signal)
Connectivity with Target Thalamus
Connectivity with Control Thalamus
A
B
C
D
E
F
Transducer
Location
L
R
R
L
(n.s.)
.005 .004 .003 .002 .001 0
p
.005 .004 .003 .002 .001 0
p

**Figure S2)** Whole brain results in MNI space using average subject (MNI-space) T1w images as a background. For all analyses, statistical maps were obtained using a fixed effects model as implemented in FSL 6.0.1, and are shown at two levels of cluster correction for multiplicity. For inhibition/reduction,a cluster-defining threshold (CDT) set at $p < 0.005$ is in blue and CDT at $p < 0.001$ is in violet. For activation/enhancement, a CDT set at $p < 0.005$ is in red and CDT at $p < 0.001$ is in yellow. **A)** Regions of significant change in BOLD signal during sonication compared to inter-sonication periods (i.e., baseline). **B)** Regions of significant BOLD change predicted by behavioral recovery. **C)** Connectivity changes observed during LIFU-on blocks compared to LIFU-off blocks (PPI) between the whole brain and the targeted thalamus. **D)** PPI changes (target thalamus) predicted by behavioral recovery. **E)** Connectivity changes observed during LIFU-on blocks compared to LIFU-off blocks (PPI) between the whole brain and the non-targeted (control) thalamus. **F)** PPI changes (non-target thalamus) predicted by behavioral recovery.

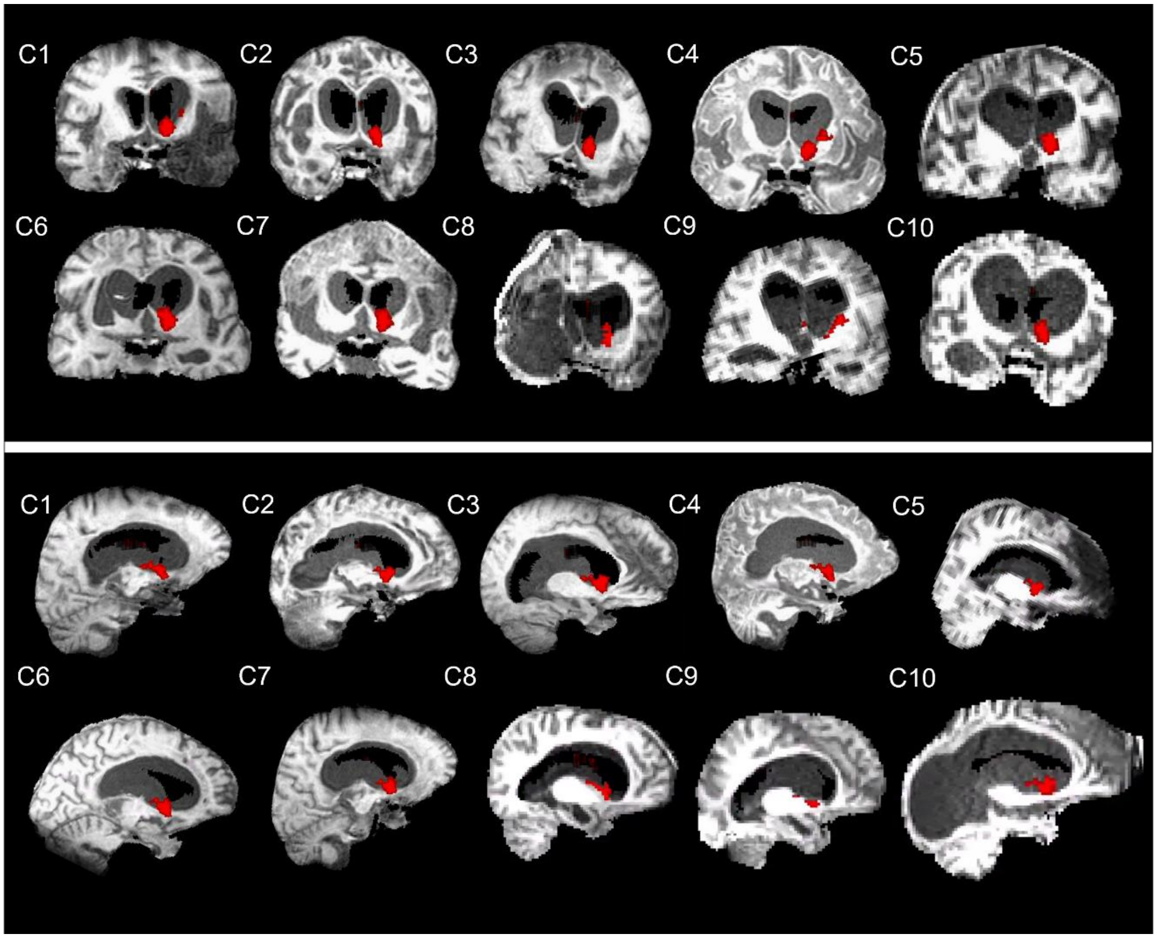


**Figure S3)** Effect of LIFU Corrleated with Recovery in Subject Space. Here, the group-wide effect seen in Figure S2 (top right) is shown in subject space for each subject in both coronal and sagital slices. Placing the location of small activations can be difficult given the idiosyncratic nature of chronic DOC brains, which should be clear from this figure. However, we believe this figure should make the reader more confident in our conclusion that, for most individuals, the activation in question appears to subsume portions of the striatum, and more specifically along the length of the caudate.

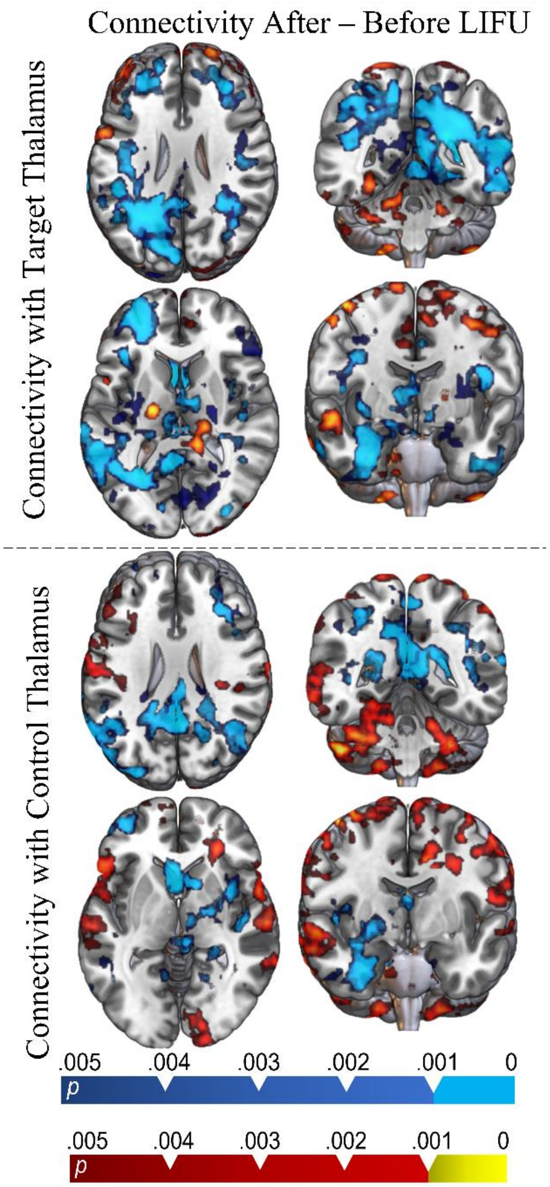


**Figure S4)** Connectivity changes with target and control thalamus following LIFU in MNI space.

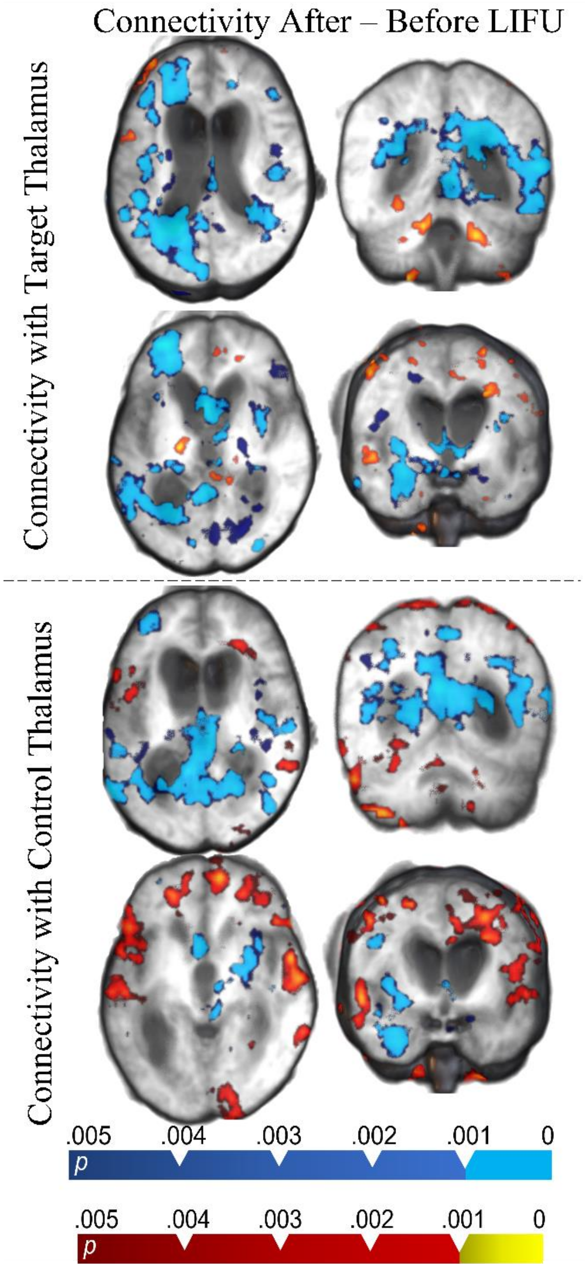


**Figure S5)** Connectivity changes with target and control thalamus following LIFU in MNI space with subject average T1 brains used as background.

### Supplementary Methods

#### MRI Sequences

Prior to transducer placement and targetting, a high-resolution T1-weighted MPRAGE (TR = 2.08, TE = 4.14, voxel size = 1x0.5x0.5mm$^3$) was acquired for later processing. Rapid (95 s) T1-weighted structural sequences (TR = 1900 ms, TE = 2.2 ms, voxel size 2 mm$^3$) were used for targetting purposes. Concurrent with LIFU administration, BOLD data was collected with a T2*-weighted Echo Planar Image sequence (TR = 2s, TE = 25ms, voxel size = 3.44x3.44x4.25mm$^3$).

#### *MRI Data Analysis: Preprocessing*

MRI data preprocessing and analysis was conducted using FSL (FMRIB Software Library v6.0.1)[1] with in-house Bash shell scripts. In addition, second level data analysis was performed using JASP. JASP Team (2019). JASP (Version 0.11.1).

In order to produce group-level functional results, data from patients who received LIFU to the right thalamus was flipped such that the right hemisphere became the left hemisphere. This inclues structural data for the purpose of co-registration. Next, preprocessing was performed including brain extraction (using optiBET [2], given its superiority in patients with DOC), spatial smoothing (using a Gaussian kernel of 5 mm full-width half-max), slice timing correction (Fourier-space time-series phase-shifting), highpass temporal filtering (Gaussian-weighted) at 0.01 Hz, and motion correction (MCFLIRT)[1,3]. With the exception of brian extraction, these were perofrmed in fsl FEAT.

Following the guidelines set by recent exploration of ideal preprocessing in DOC patients[4], head motion was mitigated by including in the statistical model a number of nuisance regressors including individual timepoints with excessive motion (i.e., spike regression [4]) derived from the output of fsl FLIRT, 24 head motion parameters, and regressors for white matter and CSF components. White matter and CSF regressors were produced by segmenting T1 images for each patient using FSL Fast (visually inspected for accuracy). Tissue segmentations for white matter and CSF were moved into functional space (for some patients, FSL epi_reg was employed while FLIRT worked better for other subjects) and binarized (and again visually inspected for accuracy). Nonlinear registration was avoided given the highly unique nature of chronic DOC patient brains (e.g., large ventricles), even when compared to acute DOC patients[5]. Time series for white matter and CSF were then extracted from functional images using fslmeants. Framewise displacements for each volume were derived from FSL MCFLIRT[1,3] and these were used to exclude unwanted volumes with a framewise displacement exceeding 0.5mm (25% of voxel width). Any functional data not exceeding 4 minutes of total time[4] were excluded from this analysis. This included only the single run from subject C5.

In order to register structural images to functional space, then, functional images to standard space, we employed a combination of FSL epi_reg, which is taylored for coregistration of subcortical regions in particular (including the LIFU target) and conventional 12 dof linear coregistration (using FSL FLIRT). Both were run for each registration and visually assessed for which was more successful. In order to register structural images to standard space, the same process was employed.

#### BOLD Data Analysis: Activity

BOLD data collected during LIFU were first analyzed employing a univariate general linear model (GLM) approach [6] including pre-whitening correction for autocorrelation (FILM). A univariate analysis was conducted using a single "task" regressor— which represented the onset time of 30s blocks of LIFU administration. Thus, here, the "baseline" condition used were the inter-sonication periods where no LIFU was applied. For each BOLD sequence, we computed 2 contrasts: LIFU > no LIFU and LIFU < no LIFU and assessed each using a Fixed Effects model given the low sample size. For patients with two LIFU exposures (all by one), results from the two runs were averaged at level two prior to third-level fixed-effects analysis. At the third level, data were cluster corrected for multiple comparisons using a cluster-level threshold of $z > 3.09$ (corrected $p < .05$). A separate level 3 analysis was conducted with cluster correction at $z > 2.57$ (corrected $p < .05$)[7]. Z-scores of 3.09 and 2.57 correspond to p-values of 0.001 and 0.05 respectively.

In order to determine if the degree of LIFU-induced modulation is associated with subsequent neurobehavioral change, an additional regressor was included in the third-level (group) analysis capturing each subject's recovery post-LIFU measured using the $\text{CRS-R}_{\text{index}}$.

#### BOLD Data Analysis: Connectivity (PPI)

In order to determine whether the connectivity of the thalamic was modulated by LIFU sonication during each 30s sonication block, we performed a psycho-physiologiocal interaction (PPI) analysis, a technique designed to detect changes in connectivity between a seed region and the rest of as a function of the onset and offset of a psychological task [8]. In this case, we were interested in changes in connectivity which occurred as a function of the onset and offset of LIFU and so our "psychological" regressor reflects those. Given the difficulty of segmenting the thalamus in these patinets (given highly unique neuroanatomy related to their condition), a left and right thalamic seed was created for each subject by manually selecting the location of each thalami and generated a 5mm sphere in each location. These masks were then moved into functional space using the same transformations used to do so for the whole-brain analysis. The time series of thalamic BOLD was extracted from each functional run using a mask for each thalamus with fslmeants. The PPI was estimated for each patient separately and aggregated at the group level with the same procedure as outlined above for the full-brain analysis.

In order to determine if the results of this PPI analysis covaried with behavioral recovery, we included, in the group analysis, a regressor describing each subject's behavioral change post-LIFU as measured using the $\text{CRS-R}_{\text{index}}$.

#### BOLD Data Analysis: Connectivity (Pre – Post)

For most subjects, resting state funcional BOLD data was collected both prior to and following ultasound application in order to determine if there were changes in connectivity between our targetted region and the rest of the brain. We were unable to collect this type of data from subject C8 and C10 because time limitations following LIFU administration and so these subjects were excluded from this analysis. Fistly, funcitonal data had been flipped in subjects that received ultrasound to the right thalamus such that final z-statstic maps would line up in terms of which hemisphere was impacted. Preprocessing of this data was carried out using an identical method as that described for the online BOLD. In this case, no data were cut from this analysis during preprocessing. Following this, level 1 analysis involved only nussiance regressors and one describing the connectivity of both the target and non-target thalami for each subject and for each session. Capturing the pattern of thalamic connectivity followed an identical procedure as that decribed in the determination of thalamic connectivity in our online PPI analysis. This produced subject specific maps for connectivity between each thalamus before and after LIFU for two sesssions per subject. Level two analysis agregated each run for each subject and subtracted connectivity after LIFU from connectivity before LIFU, which can be represented by the simple formula: MEAN ((POST_LIFU1 - PRE_LIFU1) , (POST_LIFU2 – PRE_LIFU2)). Finally, level 3 aggregated these results among subjects using a Fixed Effects model appropriate for a low number of subjects; cluster correction was used to correct for multiple comparisons using both a threshold of $z > 3.09$ ($p < 0.05$) and $z > 2.56$ ($p < 0.05$) in separate level 3 analyses. Z-scores of 3.09 and 2.57 correspond to p-values of 0.001 and 0.05 respectively.

#### Thalamic ROI Effect

In order to determine if a change in BOLD signal was observed in the thalamus itself during LIFU, thalamic ROIs created using fslmeants (as expalained in more detail above) were used to extract the mean z-score representing the change in BOLD observed during LIFU within each thalamus, each run, and within each patient. These values were asssessed in a two-way repeated measures ANOVA. Moreover, z-scores for each thalamus for each run were averaged within subject. These values were assessed for correlation with $CRSR_{index}$ scores.
